# Classification of Psychotherapy Interventions for People with Schizophrenia: Development of the Nottingham Classification of Psychotherapies

**DOI:** 10.1101/2020.07.30.20164913

**Authors:** Matthew T. Roberts, Farhad Shokraneh, Yanli Sun, Maddie Groom, Clive E. Adams

**Author notes:** **Corresponding author** Dr. Matthew T. Roberts.

## Abstract

**Background:** Currently, there is no accepted system for the classification of psychotherapies for application within systematic reviews is timely.

**Objective:** To devise a system for classification of psychotherapy interventions – for use, initially, in systematic reviews.

**Methods:** Cochrane Schizophrenia’s Register used as the source of RCTs. After being piloted and refined at least twice, finally we applied it to all relevant trials within the register. Basic statistical data already held within the register were extracted and used to calculate the distribution of schizophrenia research by form of psychotherapy.

**Findings:** The final classification system consisted of six definable broad ‘boughs’ two of which were further subdivided into ‘branches’. The taxonomy accommodated all psychotherapy interventions described in the Register. Of the initial 1645 intervention categories within the Register, after careful recoding, 539 (33%) were psychotherapies (234 coded as ‘Thought/Action’ (cognitive & behavioural) - 1495 studies; 135 ‘Cognitive Functioning’ - 652 studies; 113 ‘Social’ - 684 studies; 55 ‘Humanistic’ - 272 studies; 23 ‘Psychoanalytic/dynamic’ - 40 studies; and 63 ‘Other’ - 387 studies).

For people with schizophrenia, across categories, the average size of psychotherapy trial is small (107) but there are notable and important exceptions.

**Conclusion:** We reported a practical method for categorising psychotherapy interventions in evaluative studies with applications beyond schizophrenia. A move towards consensus on the classification and reporting of psychotherapies is needed.

**Clinical Implications:** This classification can help the clinicians, clinical practice guideline developers, and evidence synthesis experts to recognise and compare the interventions from same or different classes.

**Summary Box:** *What is already known about this subject?:*

- Effective classification of medical interventions is a perquisite for their effective identification, detection, and grouping. This in turn is essential for comprehensive identification of randomised control trials (RCTs) for inclusion in systematic reviews.
- A vast range of psychological therapies for schizophrenia exist, however there is a great degree of heterogeneity in their methods, and little consistency in their nomenclature.
- Classification of interventions for schizophrenia exists for pharmacological therapies. However only limited attempts have been made to develop such a classification for psychotherapies, and no literature-based classifications have been attempted for use in research.

*What are the new findings?:*

- The vast majority of psychotherapy interventions for schizophrenia can be consistently and systematically assigned to five broad categories: Thought/Action, Cognitive Functioning, Social, Humanistic, and Psychoanalytic/Psychodynamic. A small minority of emerging or unique psychotherapy interventions do not fit into any of these five categories.
- Using the same classification system these categories can in turn be subdivided into branches, allowing similar forms of psychotherapy to identified with greater detail, and allowing systematic reviews of greater specificity to be conducted.
- This classification was applied to Cochrane Schizophrenia’s comprehensive register of schizophrenia RCTs. It was demonstrated to be an effective method for identifying and grouping different schizophrenia psychotherapy RCTs for the purposes of conducting systematic reviews.
- The mean size of schizophrenia psychotherapy RCTs is approximately one hundred participants, consistent across different categories of psychotherapies. Thought/Action interventions – such as cognitive behavioural therapies – account for the largest proportion of schizophrenia psychotherapy RCTs. Only a small minority of schizophrenia psychotherapy RCTs investigate humanistic and psychoanalytic/psychodynamic therapies.

*How might it impact on clinical practice in the foreseeable future?:*

- The classification system we have developed can be used for the accurate identification and grouping of different types of psychotherapies. This will allow more comprehensive, accurate, and specific systematic reviews to be conducted – in turn producing better quality evidence on the effectiveness of different forms of psychotherapy for schizophrenia.
- The classification system also has applications beyond research – and likely beyond schizophrenia – including providing a framework for laypersons and clinicians to better understand and recognise different forms of psychotherapy. It also provides a contribution, and an impetus, towards improving consensus around common language and classification of psychotherapies.
- The data on study size and distribution by category of psychotherapy – which we have produced by applying our classification system to Cochrane Schizophrenia’s comprehensive register of schizophrenia RCTs – may illuminate avenues for future research into schizophrenia psychotherapy, and identify areas in which RCTs in this area can be improved.

## Background

Accurate and systematic classification of medical interventions is integral to the practice of evidence-based medicine. Those compiling treatment guidelines often used randomised controlled trials (RCTs) as building blocks within systematic reviews (OCEBM Levels of Evidence Working Group, 2011). Comprehensive identification of RCTs is particularly important to ensure all relevant data are considered and random error and systematic bias minimised in the eventual syntheses (Adams & Gelder, 1994), (Higgins, et al., 2019). In addition, the development of a practical system of classification of psychotherapies would open novel avenues of research previously made difficult because of confusions of nomenclature.

Effective classification of medical interventions is a prerequisite for their accurate detection, description and grouping. Otherwise, similar treatments have to be identified by the diverse names assigned to them (Lokker, McKibbon, Colquhoun, & Hempel, 2015). Such classifications now exist for medications (Shokraneh, 2020), but the dynamic nature of the field of psychotherapy, as well as the limited regulation in naming/description of interventions, presents challenges to production of a classification in this area.

## Objective

To devise a practical system for classification of psychotherapy interventions for use, initially, in accurate identification of studies for relevant systematic reviews

As a secondary objective, to employ the new system to help describe frequencies and size of studies of all published schizophrenia psychotherapy RCTs

## Methods

We attempted to arrive at a working definition of ‘psychotherapy’, sought classification systems of the past to use or adapt, piloted the initial attempt at a purpose-built classification on all relevant schizophrenia psychotherapy trials, and modified the system in line with this experience.

### Definition

Existing definitions, sourced from past classifications (Table 1), were considered and discussed with an academic psychologist (MG). These definitions are often intentionally broad and not formulated with classification in mind. In the definition we formulated for this work we aimed to capture the fundamentals of psychotherapy whilst creating something understandable, practical and systematic.

**Table 1.**
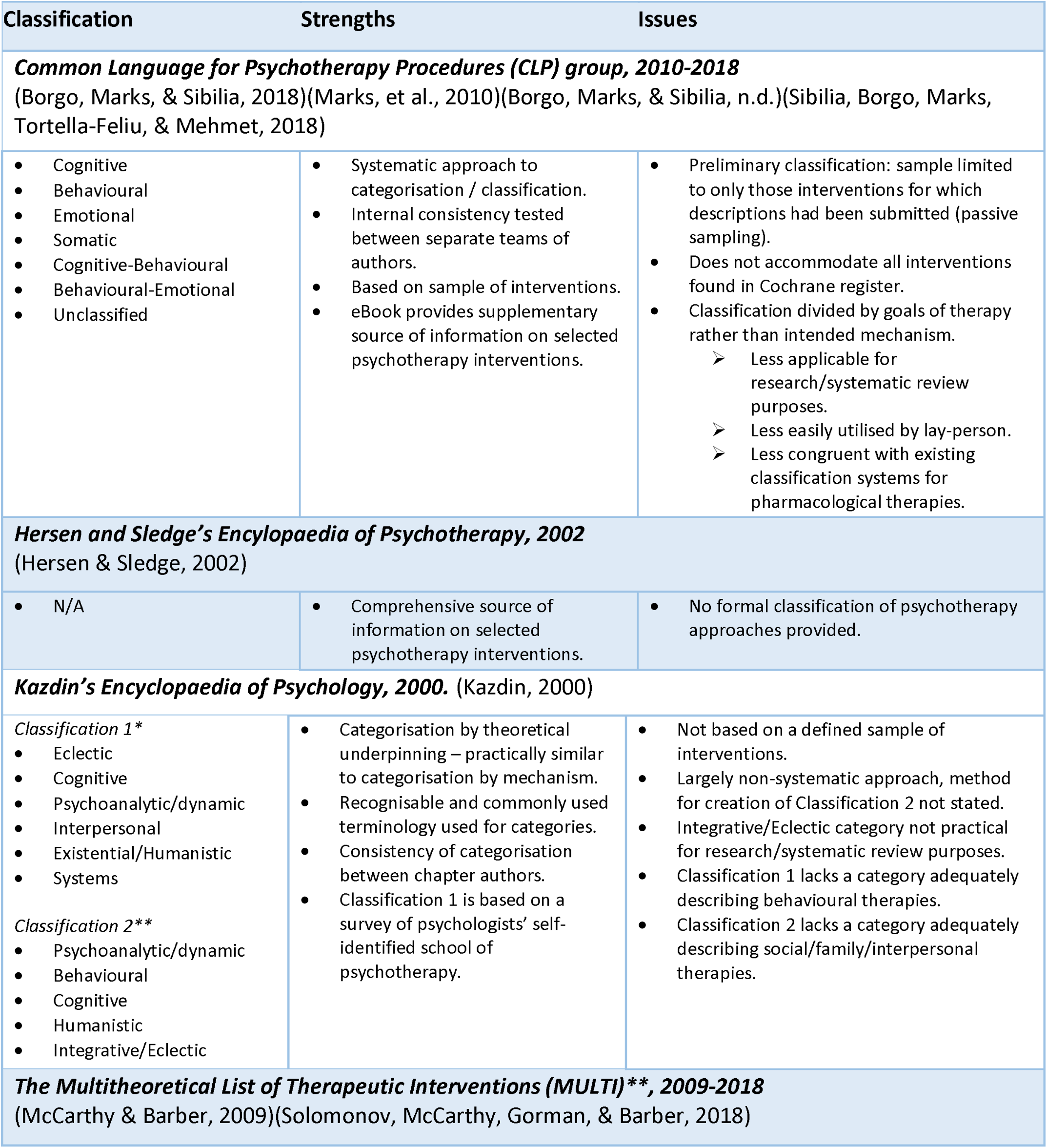
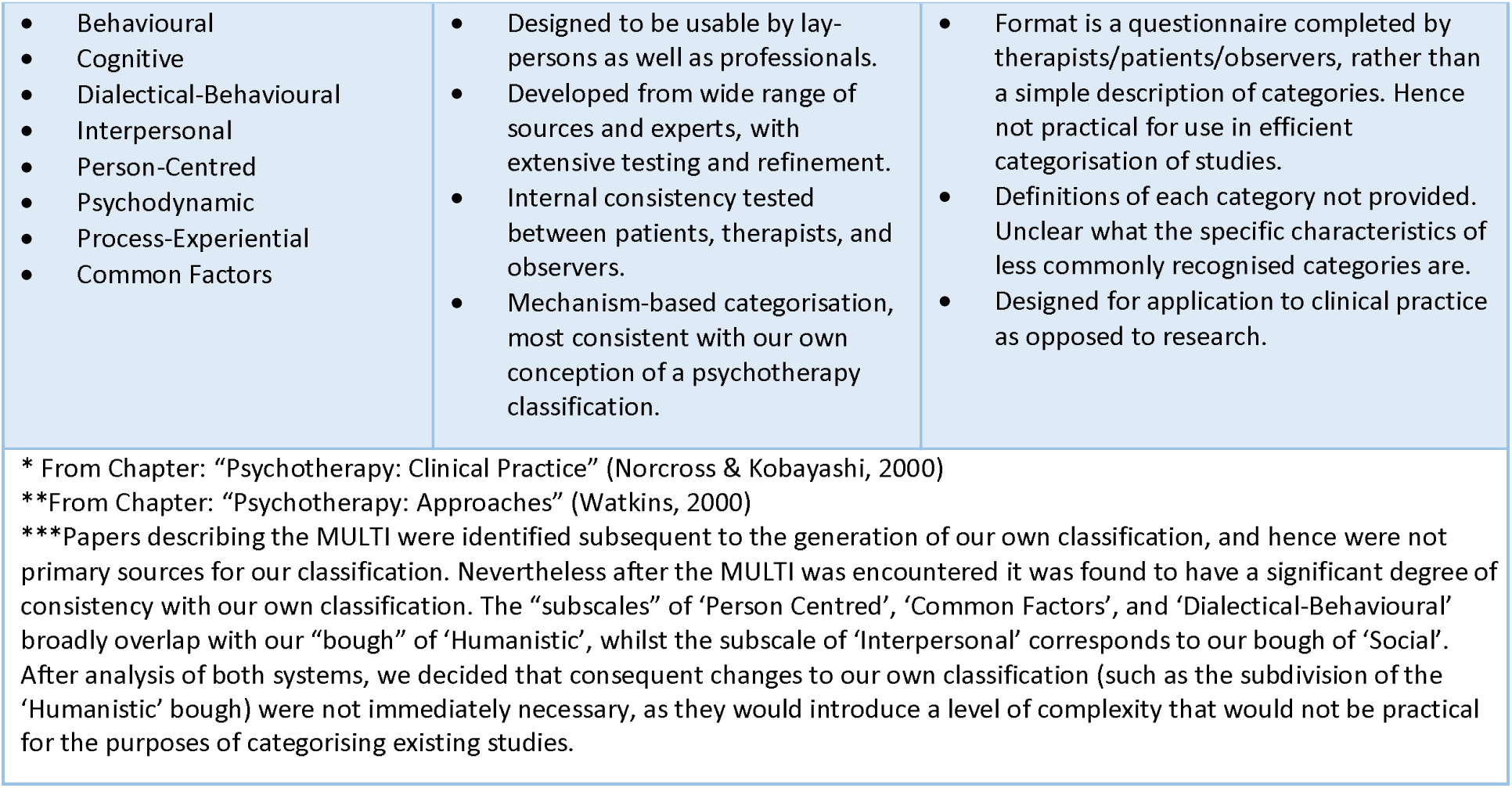
Characteristics of Previous Categorisation Attempts

**Psychotherapy – a working definition**

A treatment whose primary purpose is the improvement or prevention of direct psychological and mental sequelae of mental illness/psychopathology which:

1. Is intended to work primarily through interaction with the recipient’s mind, AND
2. Involves the active interaction of the recipient with the treatment, AND
3. Does not involve the external administration of a substance or physical effect to the recipient as its primary mechanism.

### Previous Attempts at Classification

No widely accepted systematic approach to the classification of psychotherapy interventions exists. Prominent forms of psychotherapy, such as Cognitive Behavioural Therapy (CBT), are widely recognised and grouped together through various informal groupings (Miller, 2019). However, the array of psychotherapies is vast and growing as practitioners develop novel methods. Many of these developments have names that imply overlap with past methods. However, despite similarity in nomenclature, many use different methods or theoretical underpinnings.

We sought past attempts at classification by bibliographic database searches and tabulated these. Table 1 is not a comprehensive list, but rather, outlines four overlapping but distinct perspectives we found particularly valuable. Unsurprisingly, we could not directly implement any of the previous attempts. None were designed with the specific purpose of assisting trial identification for systematic reviews.

### Source of data: Cochrane Schizophrenia’s Study-Based Register of Trials

This register – started nearly 30 years ago to facilitate the systematic review process (Adams & Gelder, 1994) – is now supported by NIHR-UK. It contains every report of every relevant randomised trial but, within it, a single *study* record is linked [related] to all relevant reports of that trial to help avoid multiple counting (Shokraneh & Adams, 2017). Creation of unique study records affords opportunity to rationalise nomenclature – to classify. This has been the case for medications, and now metadata on medication interventions have a fully controlled language (Shokraneh, 2020). This is not the case for other treatments and, currently, FS simply records the name of these other treatments verbatim from the study report with no categorisation. It is on this list of RCT-centric psychotherapy interventions the categorisation was piloted.

### Creation of the Initial Classification

We used a methods-based approach, identified examples of each uniquely named psychotherapy from the register, and divided and categorised these by their purported method of action. Our efforts were guided by the pioneering work (outlined in Table 1), the descriptions of interventions in trials and constant consideration of our primary objective – which was to produce a practical classification to aid reliable identification of relevant studies for reviews.

### Stage 1: Piloting

Our initial classification produced six reasonably distinct boughs off the main trunk of psychotherapy. These were possible to describe to aid Information Specialists assigning new studies to a bough. Any given study could be assigned multiple boughs.

The [initial] six boughs – with working definitions – were as follows:

- **Behavioural:** “Developing different behaviours leads to change”
- **Cognitive:** “Changing thought patterns leads to change”
- **Humanistic:** “Empowering the patient leads to change”
- **Psychodynamic/Psychoanalytic:** “Understanding of the self, past experiences, and the unconscious leads to change”
- **Social:** “Changing interpersonal/intergroup relationships leads to change”
- **Other:** Psychotherapies that do not fit into any of the above categories, or contain an element which is not captured by any of the above categories.

### Stage 2: Modification

As piloting on samples of trials from the register progressed, some alterations to the classification were necessary. Principle amongst these were changes made to the Behavioural and Cognitive boughs. Firstly, it was clear that two distinct groups fit under the umbrella of *Cognitive* interventions. One focuses on cognitive *content* and the other on cognitive *functioning*. ‘Content’ relates to what a person thinks about, whereas ‘Functioning’ involves emotion recognition, memory and other domains. To address this we separated the *Cognitive* bough into two – *Cognitive Content* and *Cognitive Functioning*.

Secondly – and unsurprisingly given their shared epistemological history – there was considerable overlap between many *Behavioural* and *Cognitive Content* interventions. This was most pronounced for interventions which did not explicitly name themselves as ‘behavioural’ or ‘cognitive’, but which, in their description, clearly belonged to at least one of these boughs. Even for interventions explicitly identified with either *Behavioural* or *Cognitive* schools of thought, we found significant degrees of overlap in the actual methods used. To address this we grouped *Behavioural* and *Cognitive Content* together as sub-divisions [branches] under a bough now termed *Thought/Action*. Where possible, we attempted to separate interventions into one of the two sub-divisions – but such separation is frequently impossible.

**Final Categorisation**

**Thought/Action (T/A)**

”Developing different behaviours and thought patterns leads to change”.

**Sub-division: *Behaviours***

“T/A methods primarily orientated towards changing behaviours”.

**Sub-division: *Cognitive***

“T/A methods primarily orientated towards changing thought patterns”.

**Cognitive Functioning**

“Improving cognitive functions and skills leads to change”.

**Humanistic**

“Empowering the patient leads to change”.

**Psychodynamic/Psychoanalytic**

“Understanding of the self, past experiences, and the unconscious leads to change”.

**Social**

“Changing interpersonal/intergroup relationships leads to change”.

**Sub-division: *Family*:**

“Social methods based on altering family dynamics and relationships”.

**Sub-division: *Social Skills*:**

“Social methods based on improving ability of patients to form, maintain, and utilise social relationships and networks”.

**Sub-division: *Social Support*:**

“Social methods based on directly providing patients with supportive social relationships and networks”.

**Other**

Psychotherapies which do not fit into any of the above categories, or which contain an element not captured by any of the above categories.

(*For a more detailed description of each bough – See Appendix 1*)

### Reclassification of Intervention Categories

Having piloted and modified the classification using limited subsets of trials from within the Cochrane Schizophrenia register, we now applied the classification to the intervention categories of the whole register of randomised studies (for an explanation of the structure of the Cochrane register as relevant to this analysis – see Appendix 2). FS (Information Specialist) provided a version of the register having removed all pharmacological intervention categories and traditional Chinese medicine categories (see below) (leaving 1646 unique intervention categories). For purposes of utility, this version was then further streamlined so that each remaining intervention category appeared as a single instance (linked to a single key reference). However, a complete version of the database was preserved so that all studies, references and full text PDFs linked to a specific intervention category could be viewed if required.

Each intervention category was reviewed and, where necessary, re-coded based on the classification as outlined above.

### First pass

Initially we had to exclude traditional medicines, such as Ayurvedic and Traditional Japanese medicines. As with Traditional Chinese Medicines, which had already been excluded, these involve a physical treatment but may be accompanied by some manner of psychological approach. The descriptions of these interventions were limited, and further compromised by our understanding, rendering classification impossible. We realise that this could give our classification a bias towards a ‘Western’ paradigm.

We also excluded the ‘pure’ physical therapies as many did not fit our definition of psychotherapy. The exceptions were physical therapies with a specific clear theoretical underpinning based on one of the six bough categories (such as body psychotherapy and body awareness therapy, which have a psychodynamic/analytic underpinning).

Finally, we excluded those categories which described an “Aspect” of treatment (categories which describe a generalisable aspect of an intervention rather than the intervention itself).

### Second pass

We then considered the previous labelling of the studies. Where this closely corresponded to the new classification, we reused these intervention categories without close examination of their linked reference(s). For example, the old intervention category ‘Psychoanalytic Psychotherapy’ clearly corresponded to the new ‘Psychoanalytic/Psychodynamic’ category.

A total of 205 intervention categories were coded as psychotherapy interventions using this “semi-automatic” method. Using the same approach, a further 408 intervention categories were assessed, and judged not to correspond to a psychotherapy intervention – for example ‘Transcutaneous Vagus Nerve Stimulation’.

As this process did not involve a full examination of every single linked reference for each intervention category, there was a risk of error. To address this 10% of each of the two groups described above were randomly selected and fully examined to ensure that the process was reliable. For this manual re-checking we accepted an accuracy rate of 80% and above (Appendix 3).

### Third pass

the main coding pass. Again, each category was judged as to whether it described a psychotherapy intervention. If so, it was then coded to one or more relevant boughs. Where possible each intervention category was coded to one bough and, where possible, the ‘Other’ bough was avoided. Where intervention categories clearly corresponded to more than one group, however, this was reflected in the coding. We judged that sensitivity is preferable to specificity – for a reviewer conducting a meta-analysis using the Cochrane database to identify trials for inclusion it is far more important to minimise false negatives than false positives. Although it wastes time, the latter can be manually excluded.

Coding of each psychotherapy intervention was achieved through:

1. Examination of the single randomly selected example study; and if any doubt remained:
2. Examination of other studies linked to that intervention category in the un-streamlined database segment; and if doubt continues:
3. Consulting auxiliary sources (e.g. (Hersen & Sledge, 2002), (Borgo, Marks, & Sibilia, 2018).

### Findings

1507 intervention categories were included in the analysis, of which 894 were analysed manually, whilst 613 were analysed using the semi-automatic method. The results for the full 1507 intervention categories are presented below.

Following analysis, 539 intervention categories were judged to correspond to psychotherapy interventions. The distribution of these intervention categories, in terms of boughs, studies, and participants, are presented in Table 2. 38 intervention categories were coded as adjuncts, of which 18 were also coded as psychotherapies.236 intervention categories were only linked to references in foreign languages. Of these, 230 were in a Chinese language, and were later analysed separately by a second researcher (YS), and are included in the analysis.

**Table 2:** Distribution of Analysed Intervention Categories (Primary Analysis)

|  | Interventions<br>1507 | References | Studies | Participants* | Mean participants/study<br>(Min-Max) |
| --- | --- | --- | --- | --- | --- |
| **Psychotherapies** | **539** | **6412** | **3342** | **337291** | **107(3-7628)** |
| ***Thought/Action*** | *234* | *2722* | *1495* | *161118* | *113 (3-7628)* |
| Cognitive content | 67 | 503 | 349 | 29806 | 89 (10-2160) |
| Behavioural | 108 | 829 | 523 | 64374 | 130 (3-7628) |
| Cognitive and behavioural | 59 | 1347 | 676 | 73439 | 115 (3-3780) |
| ***Cognitive Functioning*** | *135* | *1217* | *652* | *45667* | *73 (5-750)* |
| ***Social*** | *113* | *1031* | *684* | *71199* | *113 (6-1400)* |
| Family-based | 27 | 370 | 246 | 29823 | 131 (11-750) |
| Social skills | 63 | 532 | 367 | 34014 | 101 (6-1400) |
| Social support | 27 | 219 | 133 | 15456 | 129 (8-1400) |
| ***Humanistic*** | *55* | *489* | *272* | *43035* | *162 (10-3048)* |
| ***Psychoanalytic/Psychodynamic*** | *23* | *65* | *40* | *3426* | *86 (11-563)* |
| ***Other*** | *63* | *552* | *387* | *33082* | *88 (6-615)* |
| Other only | 45 | 406 | 309 | 27013 | 89 (6-615) |
| Other and additional bough | 18 | 146 | 82 | 6276 | 79 (8-360) |
| Other and adjunct | 15 | 333 | 258 | 21094 | 83 (6-615) |
| **Adjuncts** | **38** | **446** | **334** | **29156** | **90 (6-1109)** |
| Both adjuncts and psychotherapy | 17 | 334 | 267 | 21977 | 84 (6-615) |
| **Foreign Language** | **236** | **-** | **-** | **-** | **-** |
| Not Analysed | 6 | - | - | - | - |

*\*Studies for which the number of participants was unclear or not recorded were counted as having 0 participants for the purpose of this analysis and were not included in mean participant data.*
*\*\*Unavoidable slight inaccuracies in total values when sums are taken are due to occasional overlap of reference names between separate studies, and new additions to the Cochrane Registry between the point of the coding update and the analysis of distribution.*

Several of the 894 categories were eventually coded as ‘delivery methods’. They are not psychotherapies, instead they represent the way in which a person may become exposed to a psychotherapy. For instance, ‘Assertive and Outreach Support Teams’ can deliver various forms of interventions – including psychotherapies – but this intervention is not itself a psychotherapy.

Similarly, several intervention category categories were coded as ‘adjuncts’ – meaning they do not necessarily represent a ‘school’ of psychotherapy but may be used in therapy to support any form of psychotherapy. We defined adjuncts as a method of therapy designed to facilitate/complement the delivery of a psychotherapy, and, typically, one that is not used as a stand-alone psychotherapy. However, one identically named intervention could be and adjunct in one trial, and a psychotherapy in a second study. For example, art therapy can be used in a psychoanalytic manner, or can be used as an adjunct or delivery method of other therapies. Where we felt that adjuncts may also be argued to be a form of psychotherapy in themselves, we also coded them as such. We recognise that many of these therapies are still emerging, and theoretical bases are evolving. We therefore welcome and invite feedback from practitioners of these therapies for how we can better categorise them.

**Figure 1:**
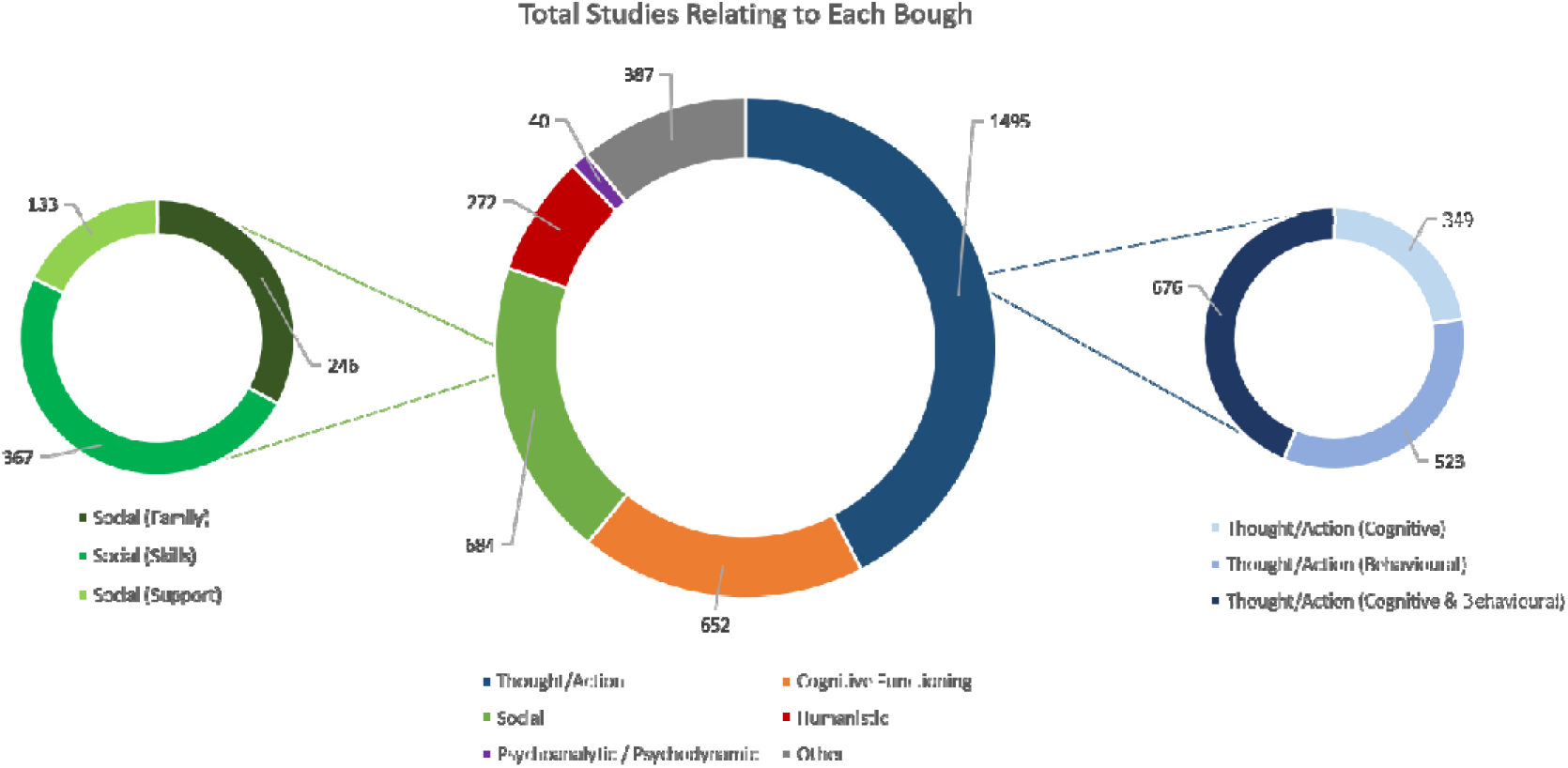
Total Studies Relating to Each Psychotherapy Bough

**Figure 2:**
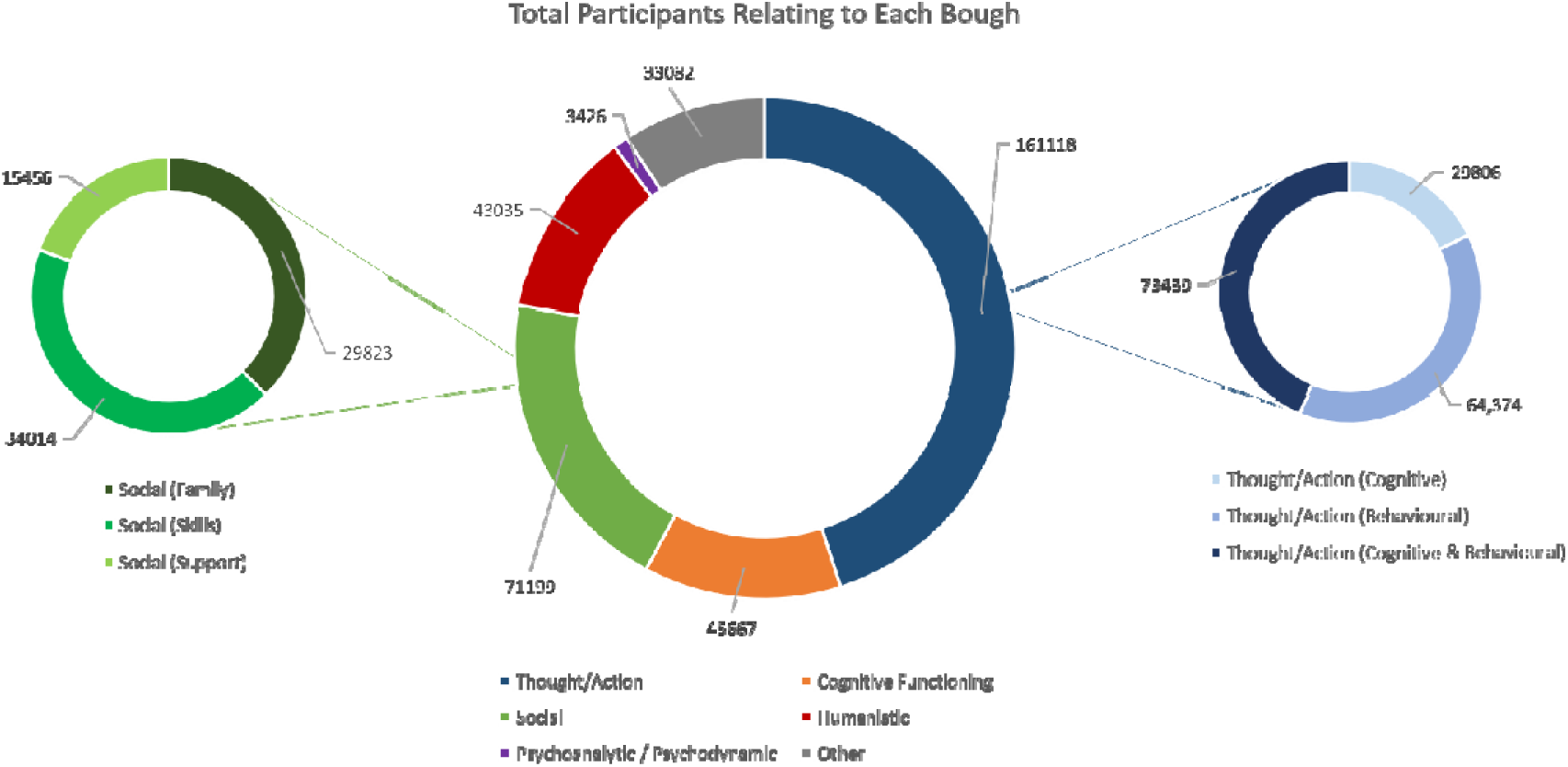
Total Participants Relating to Each Psychotherapy Bough

## Discussion

Separating the Boughs

We regard an ideal classification system for use in systematic research as one which strikes a balance between:

- Practicality/applicability: able to be quickly understood and easily applied to practical purposes.
- Recognisability: uses existing terms that are common in the psychological lexicon.
- Simplicity: avoids unnecessary introduction of layers of complexity, subdivisions, or fine-grain separating of similar categories, where these elements do not serve a practical purpose.

This classification is applicable to interventions in studies in the Cochrane Schizophrenia Register and all were able to be categorised based off a relatively rapid assessment of any given study. We were encouraged that a relatively small number of psychotherapy intervention categories needed to be coded as “Other” (over 90% of psychotherapy interventions were given a label that specifically described their ‘mechanism’). Furthermore, our classification was largely consistent with those described in past work (Hersen & Sledge, 2002)(Kazdin, 2000)(McCarthy & Barber, 2009). Whilst ours is distinct by virtue of its basis in literature rather than clinical practice, the high consistency suggests that arrival at a consensus on common language for psychotherapy classification is within reach. Achieving this goal required creation of clear, recognisable boughs, with minimal introduction of new terminology and as much reference as possible to current understanding of the sub-divisions.

In the simplest case, each psychotherapy would be assigned to a single bough and described by a single code. In practice, however, many interventions represent syntheses and combinations of theories and techniques and it would be too reductive to describe them as belonging to a single bough of the tree of psychotherapy. We, therefore, accepted the need for the possible assignment of an intervention to multiple boughs of psychotherapy and feel this approach is the most coherent with the reality of psychotherapeutic practice. Moving forward, there may be value in development of a weighting system allowing quantitative estimation of the degree to which a ‘multi-bough’ intervention corresponds to each branch of psychotherapy.

The boughs and branches must be dynamic. So long as researchers and practitioners provide clear theoretical and mechanistic frameworks for their approaches, the existing boughs should remain valid as novel therapies become established. However, once reliably coded, such classification lends itself to the capacity for swift reconsideration of the configuration of the whole tree – including the possibility of creation of new boughs/branches.

### Distribution of Schizophrenia Psychotherapy Studies

The average size of psychotherapy trial for people with schizophrenia is small (107), with little variation in size between different the boughs. These trials will have limited statistical power (Altman, 1991). However, the existence of several trials in each bough with sample sizes considerably larger than the mean, demonstrates the feasibility of conducting much more powerful studies. In the future, there would be benefit in more large, high quality, collaborative studies, achieving sample sizes of real power to investigate outcomes of importance to clinicians and recipients of care. It would also be of benefit to those classifying such studies.

The most researched psychotherapy in schizophrenia care is Cognitive Behavioural Therapy (CBT). Although the above issue of sample size does apply, this group of researchers have invested much effort testing the effects of these approaches. The clarity of description was good enough to make grouping these trials easy and this greatly facilitated producing the relevant systematic review. There is a comparative lack of research into other areas of psychotherapy – in particular Humanistic and Psychoanalytic therapies. Given the very modest – if any - benefit of CBT in comparison to other psychotherapies for schizophrenia (Jones, et al., 2018), this investment of research, clarity of description, ease of classification and supply, should allow researchers to move swiftly on to adaptions or other therapies that could be more effective.

The brief analyses presented are small and easy examples of what can be undertaken once confidence is increased in a clear classification system. Important and novel insights can be generated into research practice once classification is established, trusted and adhered to, preferably within a study-based register.

### Recommendations for Research

Journal editors could encourage authors of trials of psychotherapy to adhere to the TIDieR checklist for intervention description and replication (Hoffmann, et al., 2014) with particular emphasis on Items 2 and 3 (‘Why?’ and ‘What?’). This will increase consistency and clarity of their descriptions of psychotherapies. To strengthen the TIDieR checklist when specifically applied to psychotherapy researchers, authors should clearly – and wherever possible in simple, jargon-free language – identify:

- The school(s) of psychotherapy to which they feel the intervention belongs to; and
- The theoretical underpinnings of the intervention, including – if applicable – the progenitor psychotherapies from which it developed; and
- A comprehensive description of the practical steps involved in delivery of the intervention; and
- The specific change in a patient’s condition the intervention is intended to produce, and the intended mechanism by which the described practical steps might achieve this.

We feel that whilst this classification system was generated by work on our schizophrenia-specific register, it could be used in any register of psychotherapy literature. Information specialists whose role involves evaluation of psychotherapies would greatly enhance evolution of classification by adoption of one workable system.

### Next Steps

First, for Cochrane Schizophrenia, the register of studies will be updated with the new improved indexing categories and ‘live’ tested. This is not difficult as the register is in a MS Access database that can be updated with ease. This new indexing will be mapped onto Cochrane Schizophrenia’s publicly available topic tree (Cochrane Schizophrenia, 2020) and, in turn, relevant reviews attached to the ends of that tree. Live testing will then take place as new studies and references are added to the Cochrane Schizophrenia Register and reviews into the topic tree. Part of this ‘live testing’ will be the more detailed addition of trials not published in English. The register covers any language. In this paper the large numbers of studies in Mandarin have been accounted for but there remain some trials the detail of which remains inaccessible to us because of language.

Second, there is potential for further sub-division of boughs into [more] ‘branches’ (e.g. the *Family branch of Social*) and ‘twigs’ (sub-divisions of branches). Further analysis of each bough – and collaboration with researchers who have already attempted sub-divisions of boughs and branches– should allow incorporation of increasing levels of specificity into the classification. Examples might include sub-division of Cognitive Functioning interventions by their targeted modality, sub-division of *Humanistic interventions* by their different forms, and further sub-division of the Cognitive and Behavioural sub-divisions of Thought/Action interventions (Michie, et al., 2019).

## Conclusion

The Nottingham classification of psychotherapies is a practical, understandable, and comprehensive classification of the different boughs of psychotherapy. It is rooted in past classifications but, because its primary purpose was to allow for more accurate searching for systematic reviews of treatments, it has been primarily generated from scrutiny of existing literature.

Testing has begun on the classification and some insights into conduct of trials in this sub-speciality have already emerged. Most relevant trials are too small to be truly informative, necessitating systematic reviews for synthesis of all evidence. However, larger studies with power to investigate outcomes meaningful to routine care are clearly possible – although such trials are rare.

Application of this classification to Cochrane Schizophrenia’s register of trials will facilitate more sensitive and specific searches and allow further testing and evolution of the system. Adoption beyond this important but specific register would help validate its broad value and addition of useful finer-grain categories increases its sophistication.

In the longer term, it is vital that researchers, stakeholders, and professional organisations actively move towards global consensus on common language and classification of psychotherapeutic interventions.

## Data Availability

All the data have been reported in the manuscript and appendices.

## Contributorship

MTR has developed the protocol and ran the research and wrote the first draft of the manuscript and revised it.

FS has developed the register, contributed in finding existing relevant classification literature, read the manuscript and made comments and prepared the manuscript for submission.

YS contributed in refining the classification from a sample of trials and Chinese studies, double-checking the classed and read and commented on the manuscript.

MG research and commented on the classification and manuscript.

CEA suggested the idea for this research and supervised the team on running this research. He read, commented and revised the manuscript several times.

## Acknowledgements

Not applicable.

## Funding info

Not applicable.

## Ethical approval information

Not applicable. This is a literature-based study and does not include human participants or animal as subjects for experiment or observation.

## Appendices

### Appendix 1. Descriptions of each Bough

Below are detailed descriptions of each bough of psychotherapy in our classification, along with example codes for use in Registers. These are intended both to provide readers with a greater understanding of the nature of each bough, to allow other researchers and organisations to replicate our classification, as well as to support information specialists in the utilisation of this classification in registers.

***Thought/Action* {*T/A*}:***”Teaching different behaviours and thought patterns leads to change”*.

This predominant interventions in this bough are variations of Cognitive Behavioural Therapy, however it includes any interventions which are primarily focused on changing behaviours and thought patterns as a way of treating schizophrenia/mental illness.

Many interventions that aim to change behaviours do so by altering thought patterns, whilst many that aim to change thought patterns use behavioural techniques. For this reason we have combined cognitive and behavioural interventions under one category – however in order to distinguish interventions which are identifiably affiliated with one of the two areas, we have preserved the sub-divisions of **Cognitive {{COG}}** and **Behavioural {{BEH}}**. If an intervention cannot be easily assigned to just one of these sub-divisions, it should be coded as both (e.g. “Cognitive Behavioural Therapy {PSY} {T/A} {{COG}} {{BEH}}”).

However, this bough does not include interventions that focus on training cognitive skills and functions, such as emotion recognition (see below).

***Cognitive Functioning* {*CFN*}:** *“Improving cognitive functioning and skills leads to change”*.

This bough includes interventions which are focused on improving cognitive skills and functioning, as a means to address the cognitive sequelae of schizophrenia/mental illness. Typically, these interventions are referred to as ‘cognitive remediation’, but not all interventions of this kind will refer to themselves as such. Cognitive functioning and skills are those fundamental cognitive abilities which are present to some degree in physically and neurologically typical humans, but which can be impaired by schizophrenia and other mental illnesses (Smith & Kelly, 2015). Examples include facial and emotion recognition, memory, auditory perception, attention, proprioception, visuospatial awareness, and problem solving. For the purposes of this classification, we defined *Cognitive Functioning* psychotherapy interventions as those which treat deficiencies in these fundamental abilities arising due to mental illness/psychopathology, and aim to remediate these abilities to – or near to – the premorbid level of that person.

***Humanistic* {*HUM*}:***”Empowering the patient leads to change”*.

Perhaps the most heterogeneous of the boughs, ‘Humanistic’ therapies are those whose primary mechanism is the empowerment and personal development of the patient. Whilst many therapies may be empowering to some degree, humanistic therapies are distinct in that this empowerment and personal development is the active ingredient in change, rather than any focus on behaviours, relationships, or past experiences (Watkins, 2000).

#### Examples of humanistic therapies include

Client-Centred Therapy – which requires the therapist to avoid questions, diagnoses, reassurance, or judgement, and places the patient in charge of their treatment.

Gestalt Therapy – which involves the patient reflecting on their immediate thoughts and feelings in the current moment.

Existential Therapy – which involves a patient reflecting on their illness through a philosophical lens, based on existentialist philosophy. Patients are encouraged to develop understanding of their capacities, and to take personal responsibility for finding meaning in their life and their illness.

Similarly, an example of a partially humanistic therapy would be Dialectical Behavioural Therapy, which uses behavioural techniques to promote behaviour change, whilst simultaneously taking the humanistic approach of emphasising the patients’ acceptance of themselves as they are in the present moment. Hence, Dialectical Behavioural Therapy would be coded “Dialectical Behavioural Therapy {PSY} {T/A} {{BEH}} {HUM}”.

As can be seen, all of these therapies – whilst heterogeneous – share a common thread of patient empowerment and development as a means in itself to treating mental illness.

***Psychodynamic/Psychoanalytic* {*ANA*}:** *“Understanding of the self, past experiences, and the unconscious leads to change”*.

One of the oldest boughs of psychotherapy, Psychodynamic and Psychoanalytic therapies use exploration of a patients past experiences, as well as their unconscious thoughts and impulses, as a means both to understand the patients’ psychopathology, as well as to treat it.

Many interventions which do not specifically refer to themselves as psychodynamic of psychoanalytic fit into this category. Interventions which aim to treat schizophrenia/mental illness primarily by illuminating and/or addressing unconscious thoughts, feelings, and past experiences may be described as psychodynamic/psychoanalytic.

***Social* {*SOC*}:***”Changing interpersonal/intergroup relationships leads to change”*.

This bough includes a variety of interventions whose primary focus is on changing or utilising social relationships as a means of treating schizophrenia and other mental illnesses. It should be separated into three main sub-divisions.

Family {{FAM}}: Interventions which aim to treat schizophrenia/mental illness by changing inter-family relationships and dynamics. These can include interventions which are delivered to family members rather than directly to the patient, such as educating family members about support mechanisms and coping techniques.

Social Skills {{S-SK}}: Interventions which aim to treat schizophrenia/mental illness by improving patients’ abilities to develop, maintain, and utilise social relationships and networks. Interventions such as Interpersonal Psychotherapy would be included in this sub-division.

Social Support {{S-SP}}: Interventions which aim to treat schizophrenia/mental illness by directly building or providing patients’ with supportive social networks. This differs from the Social Skills sub-division in that the provision of a social network is the intervention, rather than the training of a patient to develop their own social networks. Examples include interventions which connect patients with peers or supporters, or enlist patients in activity and social groups.

***Other* {*OTH*}:** Psychotherapies that do not fit into any of the above categories, or containing an element which is not captured by any of the above categories.

“Other” is a broad bough that exists by necessity to describe those interventions which do not fit into any of the other boughs, but which are unusual enough not to justify the creation of a new bough. Coding interventions to this bough should be avoided where possible, but there are three main circumstances in which it may be necessary:

1. Where a previously undescribed intervention is clearly a psychotherapy, but not enough information is provided on it to assess what is mechanism is, therefore it cannot be assigned to any of the other boughs.
2. Where elements of a psychotherapy clearly correspond to a non-Other bough of psychotherapy, but other elements of the same psychotherapy are not adequately described by any of the non-Other boughs. In these instances, the intervention category should be coded as both Other {OTH} and the non-Other bough in question (e.g. “Life Coaching Program {PSY} {T/A} {{BEH}} {OTH}”)
3. Where the entirety of the intervention, whilst a psychotherapy, does not correspond to any of the non-Other boughs of psychotherapy (e.g. “Thought Discontinuation Technique (Distraction) {PSY} {OTH}”). Care should be taken with this category to take note of emerging trends – over time, with the development of new psychotherapy techniques, an entirely new bough may be justified.

### Appendix 2. How the Cochrane Register Works

The Cochrane Schizophrenia Group register is essentially a database – designed to be comprehensive – of all Schizophrenia RCTs conducted.

Each **Study** (or “Trial”) might have several unique **References** – these could be journal articles, study registry entries, conference proceedings, or any other form of reference.

Each **Reference** is linked to one or more **Intervention Categories**.

An **Intervention Category** is a description of an element of that **Reference**, and by extension its **Study**. A single **Intervention Category** may be linked to multiple different **References** corresponding to multiple different **Studies**.

*For example: a study into psychoanalytic psychotherapy might be linked to an intervention category titled “Psychoanalytic Psychotherapy”, as well as potentially several other intervention categories describing other aspects of the study*.

Each **Intervention Category** is comprised of its title and one or more **Codes** – these codes allow intervention categories to be grouped together by various commonalities, thus allowing the database to be searched such that one or more of these groups can be isolated (along with all of their linked **References**).

*For example: the full entry for “Psychoanalytic Psychotherapy” might appear as follows: “Psychoanalytic Psychotherapy* {*PSY*} {*ANA*}*”. In this example* {*PSY*} *groups this intervention category with all other psychotherapies, whilst* {*ANA*} *groups it with all Psychoanalytic and Psychodynamic therapies*.

Prior to the undertaking of this analysis there was no systematic method for (a) assigning psychotherapy interventions – in the form of references to studies of psychotherapy interventions – to intervention categories, and (b) for assigning codes to intervention categories. The construction of a new systematic classification of psychotherapy interventions described in the main text allowed all intervention categories for psychotherapy interventions to be recoded according to the “bough” of psychotherapy they correspond to.

With this completed, each reference can be reviewed and re-linked to intervention categories according to the systematic approach described in Appendix 3.

### Appendix 3. Cross Check Method & Findings

#### Method

The cross-checking process – for intervention categories which had been semi-automatically coded based on the title assigned previously by an information specialist – involved two populations: those categorised as psychotherapies and those categorised as not psychotherapies.

For each population, a 10% sample was randomly selected by assigning each intervention category an integer number ascending in alphabetical order of the categories, then using a random number generator (https://www.random.org/) to select a quantity of intervention categories equivalent to 10% (rounded up) of the total for each population.

Where any of the categories selected by this process only had foreign-language references, they were discounted and the random number generator rolled again to generate a replacement intervention category to be sampled.

Each intervention category selected by this process was then manually examined, with its linked references checked to establish whether the semi-automatic coding was accurate. This was recorded, and any inaccuracies were also amended in the main database, along with any other corrections that were inferred to be necessary during the cross-check.

Prior to the undertaking of the cross-check, it was decided that an accuracy rate of 80% - meaning that that in 80% or greater of cross-checked intervention categories for each population, the coding established by manual examination corresponded with that given during the semi-automatic stage – would be accepted. If the accuracy rate for a population fell below that number, all semi-automatically coded intervention categories would be manually re-examined and re-coded.

#### Findings

##### Psychotherapies

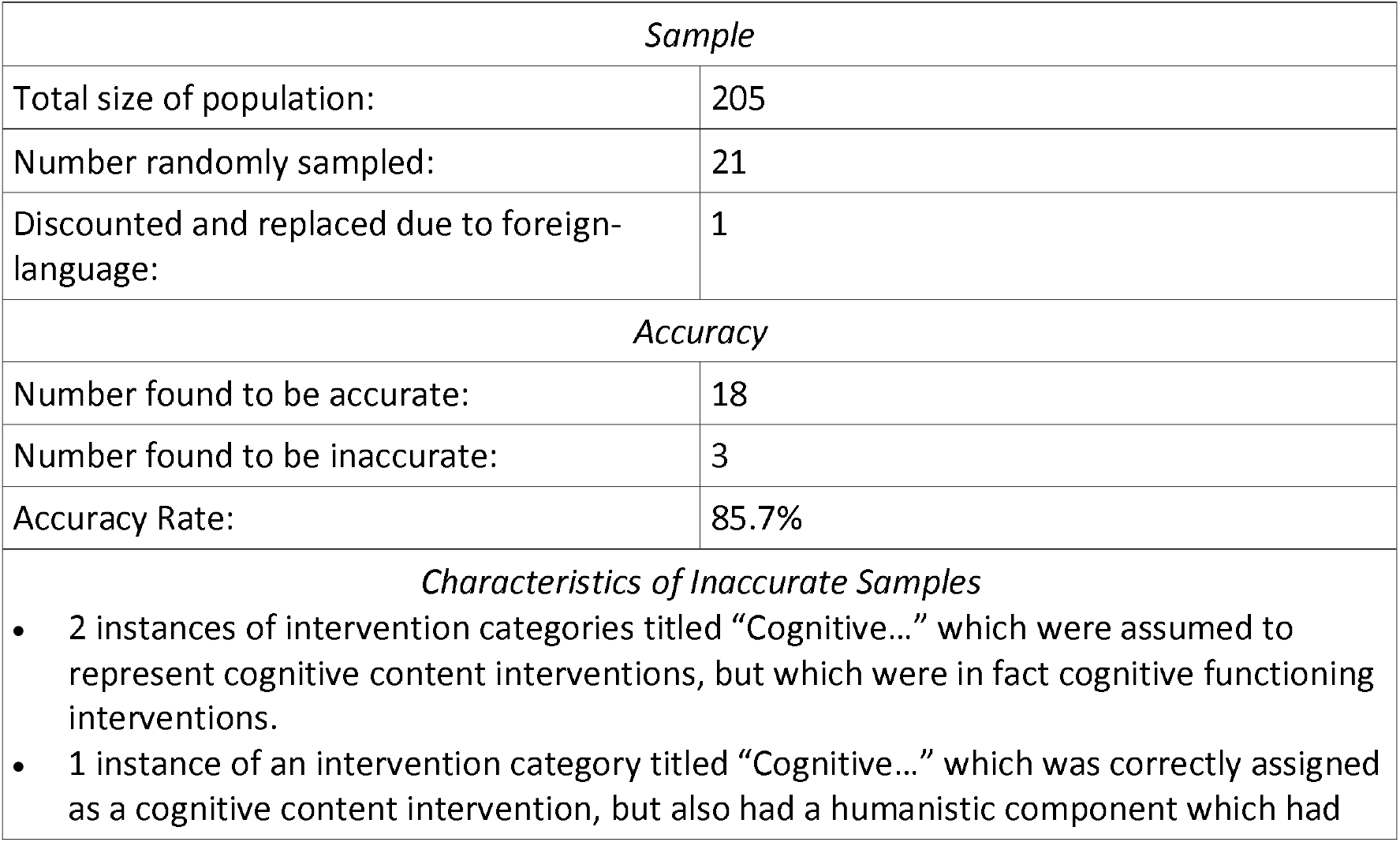
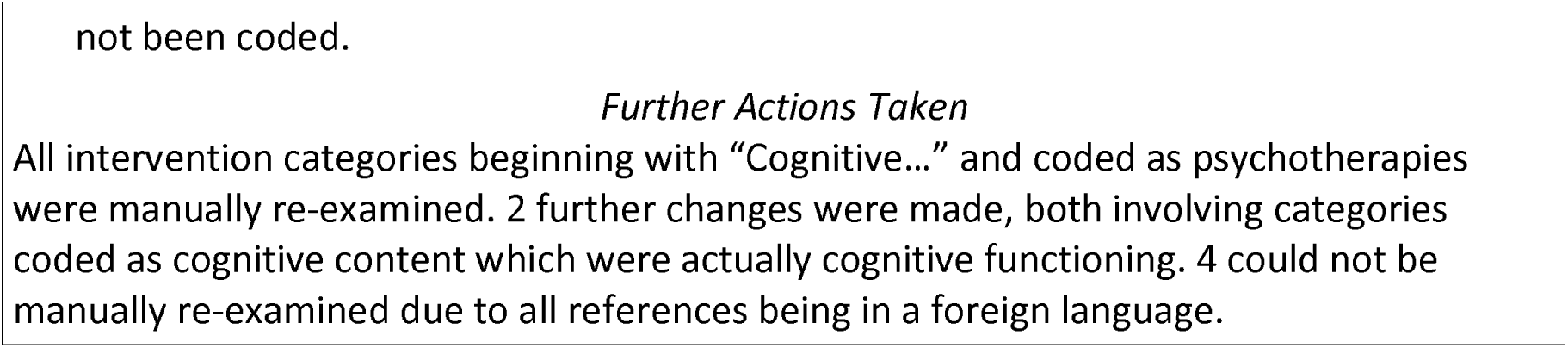

| *Sample* |  |
| --- | --- |
| Total size of population: | 205 |
| Number randomly sampled: | 21 |
| Discounted and replaced due to foreign-language: | 1 |
| *Accuracy* |  |
| Number found to be accurate: | 18 |
| Number found to be inaccurate: | 3 |
| Accuracy Rate: | 85.7% |
| *Characteristics of Inaccurate Samples* |  |
| • 2 instances of intervention categories titled “Cognitive...” which were assumed to represent cognitive content interventions, but which were in fact cognitive functioning interventions.<br>• 1 instance of an intervention category titled “Cognitive...” which was correctly assigned as a cognitive content intervention, but also had a humanistic component which had |  |

|  |
| --- |
| not been coded. |
| *Further Actions Taken*<br>All intervention categories beginning with “Cognitive...” and coded as psychotherapies were manually re-examined. 2 further changes were made, both involving categories coded as cognitive content which were actually cognitive functioning. 4 could not be manually re-examined due to all references being in a foreign language. |

##### Not Psychotherapies

| *Sample* |  |
| --- | --- |
| Total size of population: | 408 |
| Number randomly sampled: | 41 |
| Discounted and replaced due to foreign-language: | 3 |
| *Accuracy* |  |
| Number found to be accurate: | 39 |
| Number found to be inaccurate: | 2 |
| Accuracy Rate: | 95.1% |
| *Characteristics of Inaccurate Samples*<br>• 1 instance of a “Vocational Training” intervention category which corresponded to an intervention which was designed to improve cognitive functioning.<br>• 1 instance of a “Game (Visuospatial Puzzle)” intervention category which under examination fit our definition of a psychotherapy, coded as “Cognitive Functioning” and “Other”. |  |
| *Further Actions Taken*<br>All intervention categories involving vocational counselling-related interventions and not coded as psychotherapies were manually re-examined – no further changes were necessary.<br>Additionally, two further “Game” intervention categories not previously categorised as psychotherapies were manually re-examined and recoded as psychotherapies. |  |

